# Mendelian randomization study of diabetes and dementia in the Million Veteran Program

**DOI:** 10.1101/2023.03.07.23286526

**Authors:** Elizabeth M Litkowski, Mark W Logue, Rui Zhang, Brian R Charest, Ethan M Lange, John E Hokanson, Julie A Lynch, Marijana Vujkovic, Lawrence S Phillips, Richard L Hauger, Leslie A Lange, Sridharan Raghavan, VA Million Veteran Program (MVP)

## Abstract

**INTRODUCTION:** Diabetes and dementia are diseases of high healthcare burden worldwide. Individuals with diabetes have 1.4 to 2.2 times higher risk of dementia. Our objective was to evaluate evidence of causality between these two common diseases.

**METHODS:** We conducted a one-sample Mendelian randomization (MR) analysis in the U.S. Department of Veterans Affairs Million Veteran program. The study included 334,672 participants ≥65 years of age with type 2 diabetes and dementia case-control status and genotype data.

**RESULTS:** For each standard deviation increase in genetically-predicted diabetes, we found increased odds of three dementia diagnoses in non-Hispanic White participants (all-cause: OR=1.07[1.05-1.08],*P*=3.40E-18; vascular: OR=1.11[1.07-1.15],*P*=3.63E-09, Alzheimer’s: OR=1.06[1.02-1.09],*P*=6.84E-04) and non-Hispanic Black participants (all-cause: OR=1.06[1.02-1.10],*P*=3.66E-03, vascular: OR=1.11[1.04-1.19],*P*=2.20E-03, Alzheimer’s: OR=1.12 [1.02-1.23],*P*=1.60E-02) but not in Hispanic participants (all *P*>.05).

**DISCUSSION:** We found evidence of causality between diabetes and dementia using a one-sample MR study, with access to individual level data, overcoming limitations of prior studies utilizing two-sample MR techniques.

## 1.0 Background

Diabetes and dementia each impose a high healthcare burden on the U.S. population. As of March 2020, the CDC estimated diabetes prevalence at 14.7%[1] of the adult U.S. population. Separately, a 2019 study reported that 11.5% of adults at least 65 years of age had a diagnosis of Alzheimer’s disease (AD) or related dementias (ADRD), representing 1.6% of the U.S. population, a rate projected to double by 2060.[2] Observational studies have demonstrated a clear association between diabetes and dementia.[3–7] Individuals with diabetes have a 1.4-2.2 greater relative risk of dementia than those without diabetes,[3,7] with the level of increased risk varying based on the specific dementia diagnosis evaluated.[8,9] Given the projection of 17.9% prevalence of diabetes in the U.S. by 2060,[10] it is critical to gain additional understanding regarding the relationship between diabetes and dementia. The uncertainty is whether the association is due to shared genetic susceptibility, pathophysiology that independently leads to these two common diseases, or if diabetes – which typically occurs at younger ages than dementia – triggers metabolic and/or neurological changes leading to dementia.[11–13] Our previous work in the Million Veteran Program (MVP) demonstrated that a genetic risk score (GRS) for type 2 diabetes (T2D) is associated with all-cause dementia and clinically diagnosed vascular dementia (VaD), and less strongly with clinically diagnosed AD.[14] Like prior observational studies, however, our study strictly assessed an association between T2D and all-cause, VaD, and/or AD without evaluating a causal relationship.

Mendelian randomization (MR)[15] is an analysis approach that utilizes genetics to assess evidence of causality between an exposure and an outcome. In observational studies, it is difficult to establish causality because many environmental factors can influence both the exposure and outcome. The MR methodology was developed to mitigate these limitations by using genetic variation as an instrumental variable randomized at conception, therefore making the analysis less susceptible to environmental confounders. Given the widespread availability of summary statistics from genome-wide association studies (GWAS), two sample techniques[16] have become popular options for conducting MR analysis. In two-sample MR studies, summary statistics from a GWAS of the exposure are analyzed against summary statistics from a GWAS of the outcome. These techniques are efficient but do not allow for sensitivity tests requiring individual-level data. Two-sample MR studies assessing the causal relationship of T2D and other glycemic traits with the risk of AD or reduced cognitive function have found little evidence that such a relationship exists.[5,17–20] However, the opportunity to apply similar methods to assess causal relationships with outcomes such as vascular dementia has been limited by the lack of published GWAS for this specific diagnosis.[5] One-sample MR is an alternative approach that can take advantage of individual level genotype, exposure, and outcome data in a single study, as long as the sample has enough power to conduct such analysis. Large clinical biobanks which have physician diagnoses and other clinical data are well-suited to this approach.

Our objective in this study was to conduct a one-sample MR analysis in MVP, the largest clinical biobank in the U.S., to evaluate if diabetes (our exposure) causes dementia (our outcome).

## 2.0 Methods

### 2.1 Population

We conducted our study in MVP,[21] a biobank linking clinical data from the Veterans Affairs Healthcare System (VA) with genotype data on over 650,000 racially/ethnically diverse individuals.[22] The VA Central Institutional Review Board provided approval for the study protocol in accordance with the principles of the Declaration of Helsinki.[23] We analyzed MVP participants who were ≥65 years of age at the time of the release of their genotype data, stratified by harmonized ancestry and race/ethnicity classifications (HARE)[24] of Non-Hispanic White (EUR), Non-Hispanic Black (AFR), and Hispanic (HIS). HARE classifications are determined through a self-report of race/ethnicity validated with genetic data.

### 2.2 Exposure: Instruments Using Genotype Data

We utilized the study sample (N = 334,672) of participants with a coefficient of kinship ≤ 0.088, whose imputed genotype data was sourced from Release 4 of MVP 1.0 as previously described.[14,25] Our instruments were 331 variants demonstrated to be statistically significant in a T2D genome wide association analysis from the Diabetes Meta-analysis of Trans-ethnic Association Studies (DIAMANTE) Consortium[26] with European effect sizes from Mahajan et al., 2018[27] used in our previous work.[14] We chose not to use a more recent GWAS of type 2 diabetes[23] to avoid overfitting, as detailed in our previous paper.[14] The variants were not in linkage disequilibrium (R2≤0.5),[26] and for our initial steps, we retained the rs429358 variant known to be associated with both diabetes and dementia.[14]

### 2.3 Outcome: Case-Control Definitions of Dementia

Our outcomes were three clinical diagnoses of dementia (Figure S1 in the Supplement): all-cause dementia, vascular dementia (VaD), and Alzheimer’s Disease (AD), based on International Classification of Diseases 9^th^ (ICD9) or 10^th^ (ICD10) Revision[28] diagnosis codes (Table S1 in the Supplement). In the VA electronic health record, these codes are logged by presiding physicians during routine clinical care. A case was defined as a participant with at least two ICD 9 or 10 codes corresponding to one or more outcomes, as defined by the grouping in Table S1. Note that individuals meeting the case definition of AD and VaD were included as cases for both – that is, AD and VaD were not mutually exclusive. Controls were those participants without a single dementia ICD code in VA clinical records.

### 2.4 Statistical Analysis

We compared individuals with and without prevalent T2D using chi-square tests for categorical variables and t-tests for continuous variables. Prevalent T2D cases were defined as individuals whose first diagnosis occurred prior to MVP enrollment. We used MR[15] analysis to evaluate evidence of causality between T2D and clinical diagnoses of dementia. The use of MR requires that three assumptions are satisfied (Figure 1): A. The genetic variants must serve as strong instruments for the exposure (relevance); B. the instrumental variable (IV) must be related to the outcome only through the exposure (exclusion restriction, i.e., no pleiotropy); and C. the IV must not be associated with any confounders between the exposure and outcome (exchangeability).

**Figure 1.**
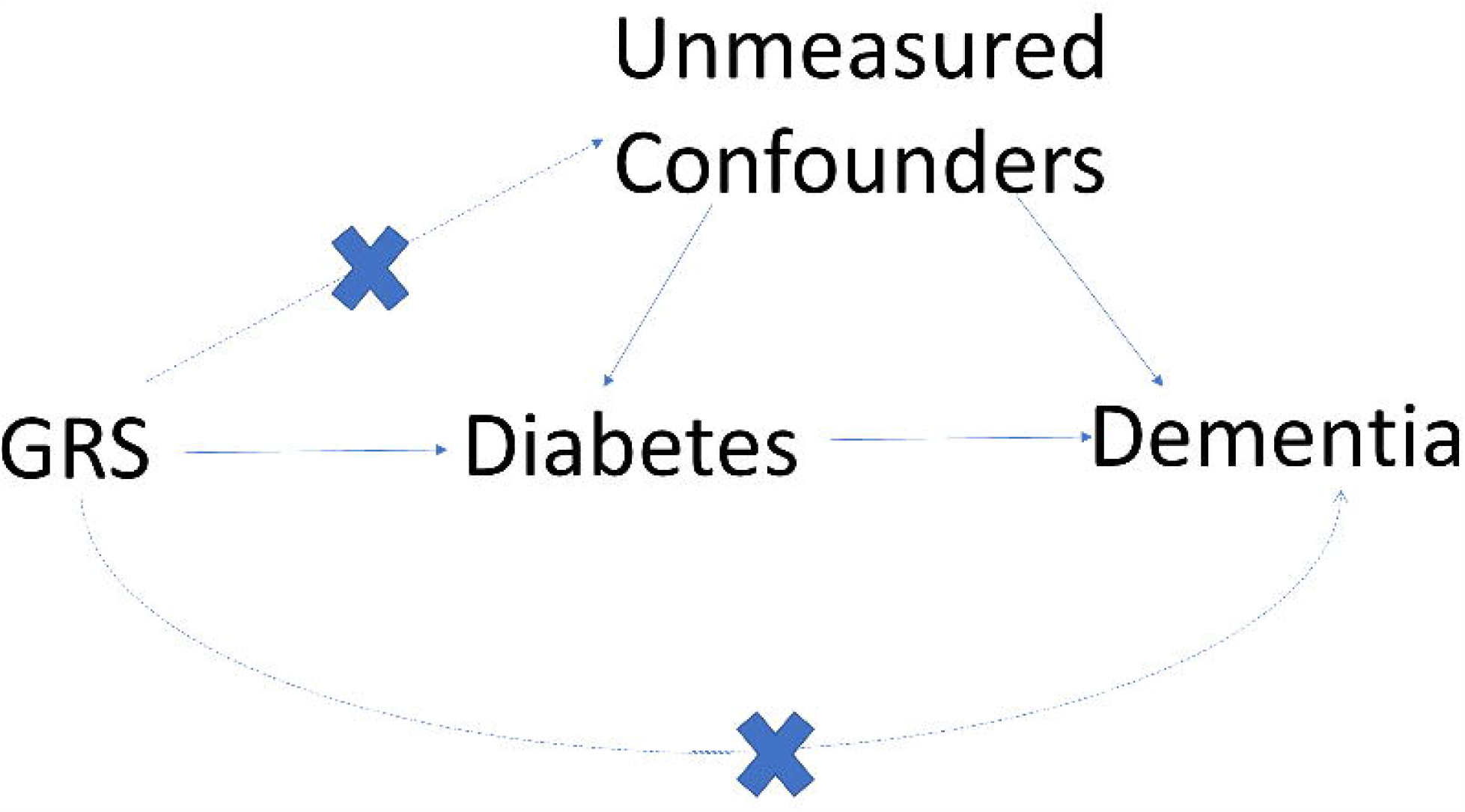
Mendelian Randomization Assumptions: The required assumptions to ensure the validity of the Mendelian Randomization Analysis Approach

### 2.5 Two Stage Least Squares Method

We used a two stage least squares (2SLS) MR[29] approach to estimate the causal associations between T2D and three clinical diagnoses of dementia. The 2SLS method employs two regression equations

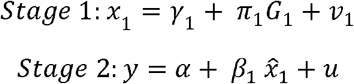

where stage 1 evaluates the genetic risk with the exposure and stage 2 estimates the prediction of the exposure with the outcome. For our genetic risk, we regressed a standardized weighted score of valid genetic instruments and effect sizes from published GWAS[30] against the log odds of T2D in the MVP biobank, adjusting for 10 genetic principal components. In stage 2, we evaluated the association of the standardized genetically predicted probability of T2D (referred to as ‘genetically predicted T2D risk’ going forward) from stage 1 with our three outcomes of dementia using logistic regression adjusting for age, and self-reported biological sex. To account for the uncertainty inherent in the first stage of the analysis, we utilized the ‘ivtools’ methodology introduced in Sjölander and Martinussen[31] to provide estimates for nonlinear models. In more precise terms, we specified the ‘ts’ estimation method in the R function ‘ivglm’ supplying the regression model output from the two stages and reported the estimates resulting from this function. The beta coefficient estimates can be interpreted as the log odds of dementia per standard deviation increase in genetically predicted T2D risk. All models were stratified by EUR, AFR, and HIS, with a significance threshold of 0.017 (0.05/3) to account for multiple testing.

### 2.6 Quartiles of Genetic Risk

To evaluate the impact of increasing genetic risk of diabetes on the three dementia diagnoses, we also assessed the difference in the odds of dementia by quartiles of diabetes genetic risk. That is, we divided the genetically predicted T2D risk from the first stage of our analysis into quartiles and evaluated the top three quartiles of risk against the reference quartile, the one with the lowest genetic risk of T2D. In this evaluation, the quartile was treated as a factor input to the second stage of the model and again, we used the ‘ivglm’ method to account for uncertainty in the first stage. Additionally, we assessed potential trends in the quartiles of progressive genetic risk using a Cochran Armitage trend test.[32,33]

### 2.7 Summarized Data and Pleiotropy Assessment

We also assessed the causal association using MR methods built to analyze effects when conducting two-sample MR. We calculated estimates using the inverse-weighted (IVW), median, and MR Egger[15] methods recommended as part of the MendelianRandomization[34] R package (v0.3.0). These methods provide effect estimates and standard errors based on summary measures of the relationship with the exposure versus the relationship with the outcome. The methods also assess evidence of pleiotropy. We additionally used MR-PRESSO[35] to test for outliers in the list of valid genetic instruments (R package version 1.0).[35] Details of these methods can be found in the Supplement (Methods S1).

### 2.8 Addressing Mendelian Randomization Assumptions

We used linear regression to evaluate the weighted instrument association strength with diabetes status by race/ethnicity population to test assumption A. An F-statistic in the linear relationship between the instrument and the exposure >10 is conventionally accepted as an indicator of a strong instrument[36]. To evaluate pleiotropy (assumption B), we used the MR-Egger method from the MendelianRandomization[15,37,38] package in R. MR-Egger constructs the best fit line in the relationship between the exposure effects and outcome effects. Pleiotropy is concluded to occur if the intercept of the slope of this relationship differs from zero. We also used MR_PRESSO[35] to check for pleiotropy. MR_PRESSO uses a leave-one out strategy to test for the impact of any single variant being considered as an instrument. If we found that any variants violated the no pleiotropy assumption, we removed those variants from the list of valid instruments for this analysis. To address assumption C, we used the recommendation by Vanderweele, et. al.,[39,40] to calculate E-Values. An E-Value puts a boundary on the potential bias introduced by unmeasured confounders. The E-Value is the effect size of an unmeasured confounder, with both the exposure and the outcome, required to explain away the exposure-outcome effect.

## 3.0 Results

We had 82,980 participants with a T2D diagnosis and 251,692 without for a sample size of 334,672 that met our study inclusion criteria (Table 1). Compared to those without T2D, those with T2D were younger (74.0 versus 74.7 years), less likely to be female (2.5% versus 3.2%), and more likely to be AFR (17.4% versus 11.4%) or HIS (7.5% versus 4.7%). All-cause dementia and VaD cases were more prevalent in those with T2D than in those without: 8.7% vs 6.7%, and 2.0% vs 1.0%, respectively. Prevalence of AD was similar between those with/without T2D (1.4% vs 1.3%).

### 3.1 Two Stage Least Squares Results

From the 331 initial variants, we identified 330 that met our criteria for valid instruments. For the remainder of our discussion, we will call the standardized weighted combination of these variants ‘GRS330’. In stage 1 of the 2SLS model, the odds of diabetes were significantly associated with GRS330 for all three HARE groups (OR of T2D per standard deviation increase in GRS330 in EUR: 1.50, *P*=4.6e-308; AFR: 1.28, *P*=5.2e-125; HIS: 1.46, *P*=3.2e-118). The standardized predicted probability of T2D risk from stage 1 (we will call this value ‘GRS330_Predicted’) was calculated for each individual and the distribution of this value differed by T2D status (Figure 2).

**Figure 2.**
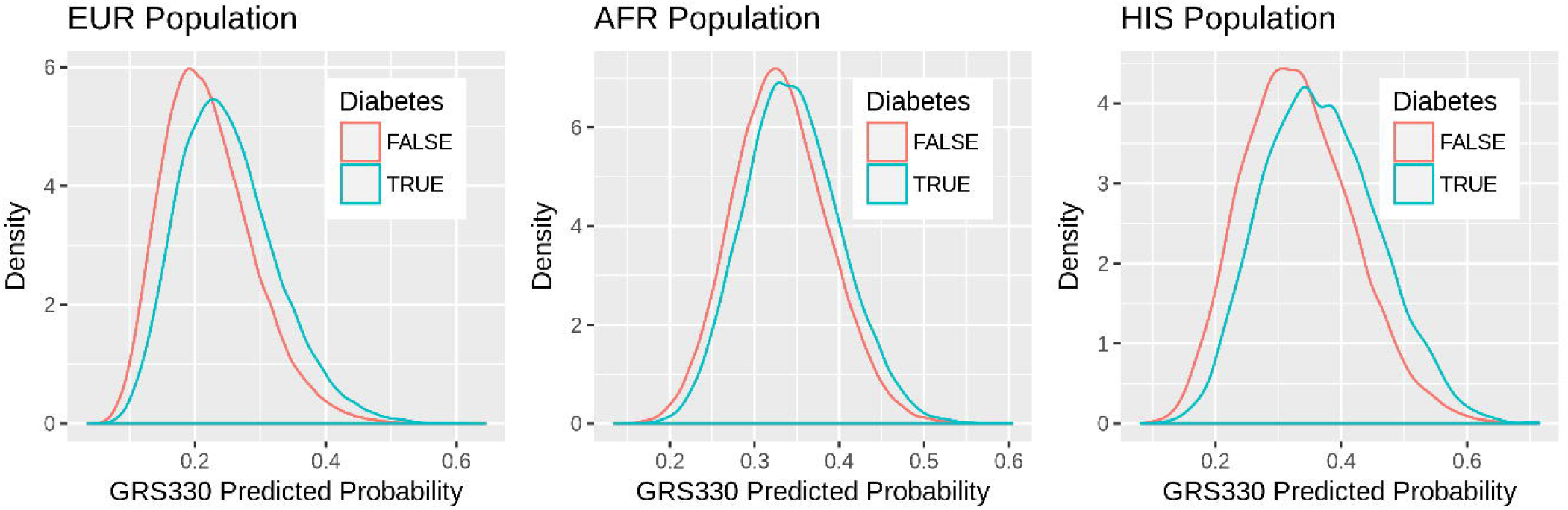
Predicted Probability: Distribution of the standardized predicted probability of GRS330 (excluding the rs429358 variant) in those with and without diabetes stratified by race/ethnicity classification.

GRS330_Predicted was associated with all dementia diagnoses (Table 2) in EUR. For each standard deviation increase in GRS330_Predicted, the odds of all-cause dementia increased by 1.07 (95% CI: 1.05-1.08, *P*=3.40E-18). We found similar results for VaD (OR=1.11, 95% CI: 1.07-1.15, *P*=3.63E-09) and AD (OR=1.06, 95% CI: 1.02-1.09, *P*=6.84E-04). In AFR, GRS_Predicted was associated (Table 2) with all-cause dementia (OR=1.06, 95% CI: 1.02-1.10, *P*=3.66E-03), VaD (OR=1.11, 95% CI: 1.04-1.19, *P*=2.20E-03) and AD (OR = 1.12, 95%CI: 1.02-1.23, *P*=1.60E-02). For HIS, there were no statistically significant associations (Table 2) between GRS330_Predicted and any dementia diagnosis (all *P*>.05).

### 3.2 Quartile Analysis Results

To demonstrate the impact of higher genetic liability for type 2 diabetes, we have also shown the risk of dementia diagnosis by quartile of GRS330_Predicted (Figure 3). In EUR, the 4^th^ quartile of GRS330_Predicted increased the odds of all dementia diagnoses as compared to the 1^st^ quartile (Figure 3a): all-cause dementia (OR=1.18, 95% CI: 1.14-1.24, *P*=3.82E-15), VaD (OR=1.33, 95% CI: 1.20-1.47, *P*=3.08E-08), and AD (OR=1.14, 95% CI: 1.03-1.25, *P*=8.13E-03). We observed similar results for the comparison of the 4^th^ quartile of genetically-predicted T2D risk to the 1^st^ in AFR (Figure 3b) for all-cause-dementia (OR=1.18, 95% CI: 1.06-1.32, *P*=3.33E-03) and VaD (OR=1.42, 95% CI: 1.16-1.74, *P*=6.63E-03), but not for AD (*P*=.07). In HIS, none of the quartile associations with dementia diagnoses were statistically significant (all *P*>.18).

**Figure 3.**
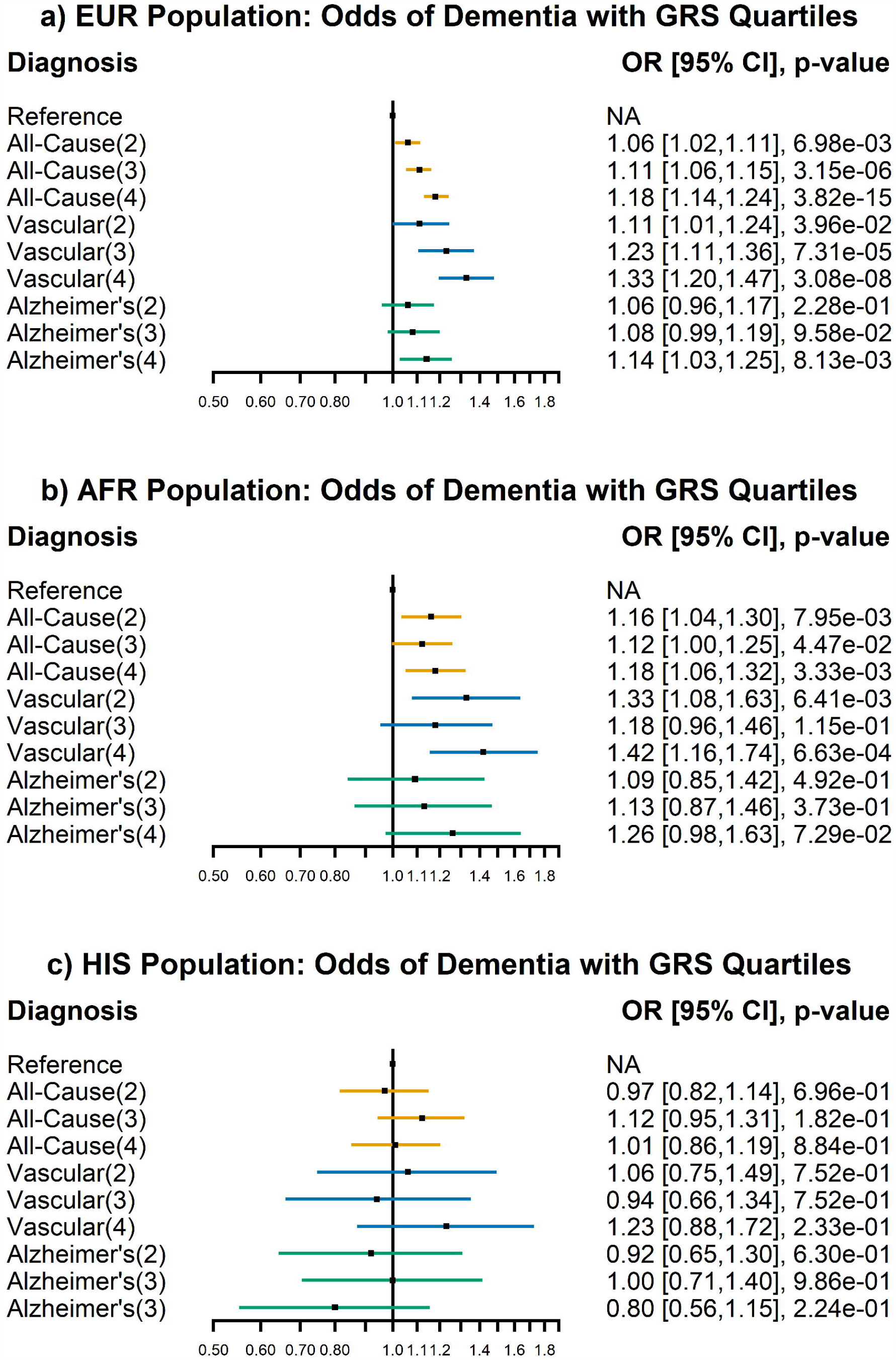
Risk Quartiles: Quartiles, listed in parentheses, of genetically predicted risk of diabetes (GRS330_Predicted) associated with increased risk of dementia diagnosis for all-cause dementia (orange), clinically diagnosed VaD (blue) and clinically diagnosed AD (green) in a) EUR: European population, b) AFR: African population, c) HIS: Hispanic population

The Cochran-Armitage[32,41] test (Table S2) for the trend in the relationship between GRS330_Predicted quartiles and dementia diagnoses was significant for all-cause dementia (*P*=3.6e-05) and VaD (*P*=3.6e-05) but not AD (*P*=.82) in EUR. The trend tests for GRS330_Predicted quartiles and dementia were not significant for any diagnosis in AFR (all *P*>.05) or in HIS.

### 3.3 Summarized Data Results

When utilizing the two sample MR approaches, we used the weighted combination of the instruments, GRS330, for the exposure. All methods for assessing the association between GRS330 and all-cause dementia in EUR provided consistent directions of effect with the primary 2SLS analyses and significant associations (Figure 4a, Table S3 in the Supplement). The results for AFR (Figure 4b) and HIS (Figure 4c) were directionally consistent with those of EUR except for the MR Egger approach in AFR (Figure 4b), though the associations were not always statistically significant (Table S3 in the Supplement). Similarly, the two sample methods provided consistent results with the primary one-sample analysis for the dementia subtypes, though not always achieving statistical significance (See Results S1 in the Supplement for full details).

**Figure 4.**
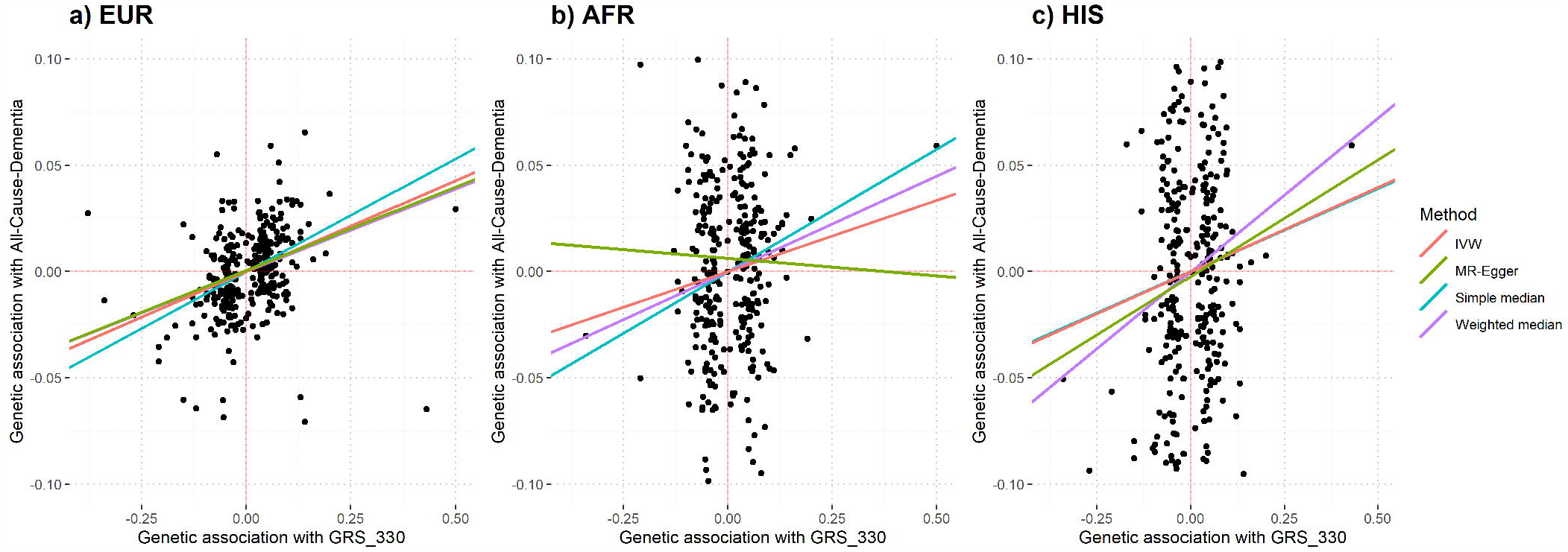
Mendelian Randomization Methods: Methods using the Mendelian Randomization package are shown here. Simple estimates have no weighting and weighted estimates use standard errors for weighting factors. a) Associations between GRS330 and all-cause dementia in EUR, b) Associations between GRS330 and all-cause dementia in AFR, and c) Associations between GRS330 and all-cause dementia in HIS.

### 3.4 Assessment of Mendelian Randomization Assumptions

Assumption A. Strong Instrument (relevance): The combination of the final 330 valid instruments (GRS330) demonstrated a significant association with diabetes. The F-statistics in the linear associations between the instruments and diabetes were 61.8 for HIS, 57.3 for AFR, and 740.4 for EUR.

Assumption B. Pleiotropy (exclusion restriction): The MR Egger test with GRS330 did not exhibit pleiotropy (*P*>0.05, Table S4 in the Supplement). As expected, however, when additionally including the rs429358 variant known to be associated with both T2D and dementia, heterogeneity was detected (Table S4 in the Supplement, all *P*<.017) except in the case of HIS with VaD. This heterogeneity was attenuated when removing rs429358 from the test (all *P*>.01). The MR_PRESSO test (Table S5 in the Supplement) characterized rs429358 as an outlier in the relationship between the exposure effect and outcome effect for all HARE groups and all clinical dementia diagnoses except HIS with VaD. Given the results of the pleiotropy and heterogeneity assessment, we excluded rs429358 and used GRS330 as our valid instrument.

Assumption C. Unmeasured Confounders (exchangeability): The sensitivity E-Value calculations were based on our previous work[14] in which we calculated odds ratios for the association of GRS330 with each clinical diagnosis of dementia stratified by HARE group. We used the risk ratio formula recommended by VanderWeele and Ding[40] for the scenario in which disease prevalence is less than 15%. As the odds ratios ranged from 1.02 to 1.15, the E-Value ranged from 1.16 to 1.57 with the E-Value of the lower bound of the 95% confidence interval ranging from 1.0 to 1.4. (Table S6 in the Supplement).

## 4.0 Discussion

We found evidence of causality between T2D and all-cause dementia as well as clinically diagnosed VaD in EUR, with similar results in AFR. The effect estimates in HIS were similar to EUR and AFR but without statistical significance. The relationship between T2D and clinically diagnosed AD had a reduced effect estimate in EUR in comparison to all-cause dementia but an increased effect estimate in AFR, indicating a need for further research to elucidate the differences by HARE group.

Given the wealth of observational data linking T2D and dementia, researchers have hypothesized a causal relationship between the two but have found little evidence to support such a conclusion. In work focused on AD as an endpoint, several studies found no evidence of a relationship with genetically predicted T2D risk: a Danish study by Thomassen et. al.,[5] the International Genomics of Alzheimer’s Project by Østergaard et. al.,[18] the Health and Retirement Study (HRS) by Walter et al.,[17] and DIAbetes Genetics Replication And Meta-analysis (DIAGRAM) and Meta-Analyses of Glucose and Insulin-related traits Consortium (MAGIC) by Pan et. al.[42] When focused on general cognitive impairment as the outcome, Ware et. al.[20] (HRS) and Garfield[19] (UK Biobank) had similar results. These results could be explained by the lack of broad genetics studies for other dementia diagnoses such as vascular dementia, as noted by Thomassen et.al.,[5]. Our study, which did identify evidence of causality, was able to assess multiple diagnoses of dementia using recently reported T2D genome-wide significant variants.

While the previous studies linking the broad spectrum of T2D with AD did not report evidence supporting causality, studies deciphering mechanisms related to diabetes have been able to offer further insight. Walter et. al.,[17] discovered that genetically-predicted insulin sensitivity was causally associated with AD in HRS. In a like manner, Pan et. al.,[42] detected a causal relationship when assessing genetic instruments for higher fasting glucose and lower HOMA-β-cell function with AD using a two-sample MR approach across the DIAGRAM and MAGIC consortia. Tschritter et al.,[12] demonstrated a cortical activity response difference to insulin infusion between carriers and non-carriers of the 972Arg variant of the *IRS-1* gene known to be associated with type 2 diabetes[43], as well as a similar response difference between lean (insulin-sensitive) and obese (insulin-resistant) individuals. These studies suggest future research directions that delve into the potential metabolic pathways through which diabetes might lead to dementia.

Previous studies assessing the relationship between T2D and AD were focused on populations with European ancestry. The MVP biobank is well powered to assess associations in an African American population. In particular, we discovered evidence of causality between T2D and clinically diagnosed VaD in AFR to have a similar magnitude of effect to EUR even though the sample size was smaller. The higher effect sizes for clinically diagnosed AD (Table 2) and the outlier effect observed with all-cause dementia (Figure 4b) in this population suggest an area for focused follow-up in future research.

We had several limitations in our study. The small number of female participants does not allow an assessment of distinctive results by biological sex. Given the small proportion of females, our results are largely driven by male participants, and so cannot be generalized to both sexes. Our outcomes were determined based on a physician’s diagnosis which may cause some amount of imprecision. Our T2D weighting structure was based on a previous study of European ancestry that might overlook important genetic architecture specific to the African American and Hispanic populations.

Despite these limitations, we conclude from our one-sample MR study in a large US biobank that there is evidence of a causal association of diabetes with dementia. Moreover, the difference in the strength of effect in the association of vascular dementia versus AD in the European population is suggestive of different mechanisms. The strength of association for both vascular dementia and AD in the African population requires further investigation. Establishing a causal association represents a first step toward examining the potential impact of the expanding prevalence of diabetes on dementia incidence and whether diabetes prevention and/or treatment can mitigate dementia risk.

## Data Availability

Summary data produced in the present study are available upon reasonable request to the authors

## Author Contributions

EML designed the study, performed the literature review, conducted the analysis, and wrote the manuscript. MWL contributed to the dementia phenotype definitions and edited the manuscript. RZ implemented the dementia phenotypes. BRC implemented the diabetes phenotype. EML(2) provided guidance on statistical methods and edited the document. JEH provided guidance on applicable epidemiology concepts and study design. JAL designed the dementia phenotype groupings. MV provided epidemiology advice on the MR study design. LSP designed the diabetes complications study. LAL provided epidemiology, study structure and writing guidance. RLH provided dementia subject matter expertise. SR offered diabetes subject matter expertise, study design and analysis guidance, and edited the manuscript. EML and SR are the guarantors of this work and, as such, had full access to all the data in this study and take responsibility for the integrity of the data and accuracy of the data analysis. The corresponding author attests that all listed authors meet authorship criteria and that no others meeting the criteria have been omitted.

Preliminary results of this study were presented as a poster: E.Litkowski,et al., Evidence of causality between type 2 diabetes and dementia in the Million Veteran Program; Abstract/Program 3896. Presented at the 71st Annual Meeting of The American Society of Human Genetics, October 18, 2021, Virtual.

## Consent Statement

MVP participants went through a counseling process before they enrolled and provided consent to have their electronic health records reviewed. The VA Central Institutional Review Board gave approval for the study protocol in accordance with the principles of the Declaration of Helsinki.

