## Supplemental Material for "Mendelian randomization study of diabetes and dementia in the Million Veteran Program"

Figure S1: Nested Levels of Dementia Outcomes, p. 1

Table S1: Dementia Subtypes and Corresponding ICD 9 and 10 Codes, p. 2

Methods S1: Two-Sample MR Approaches, p. 3

Table S2. Cochran-Armitage trend test, p. 3

Table S3. MR estimates for all dementia diagnoses by HARE group, p. 4

Results S1. Two-Sample MR Results, p. 7

Figure S2. Methods using the Mendelian Randomization package, p. 8

Table S4. Pleiotropy assumption, p. 9

Table S5. MR_Presso Results, p. 9

Table S6. Sensitivity E-value, p. 10

Table S7. Table of results when adjusting for BMI, p. 10

References, p. 12

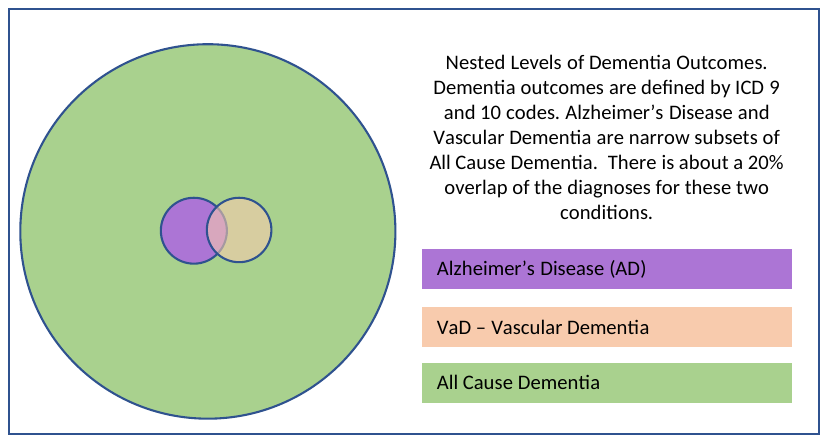

**Figure S1:** Nested Levels of Dementia Outcomes. NOTE: Dementia outcomes are defined by ICD 9 and 10 codes. Clinically diagnosed Alzheimer’s disease and vascular dementia are narrow subsets of all-cause-dementia. There is about a 20% overlap of the diagnoses for these two conditions.

**Table S1:** Dementia Subtypes and Corresponding ICD 9 and 10 Codes. The full set of dementia codes are provided to show the context of possible diagnoses. Our analysis focused on clinically diagnosed AD, VaD, and all-cause-dementia which encompasses the entire table.

| **Cognitive Disorder** | **ICD-9-CM Diagnostic Code** | **ICD-10-CM Diagnostic Code** |
| --- | --- | --- |
| ***Alzheimer’s Disease AD*** | | |
|  | **331.0** Alzheimer’s Disease, Excludes early onset AD ICD 10: G30.0 – AD+, Excludes subjects with an ICD code for AD prior to age 65 | **G30.1** Alzheimer’s Disease, with late onset, G30.8 Other Alzheimer’s Disease, G30.9, Alzheimer’s Disease, unspecified |
| ***AD+ = AD + Non-specific Dementia Listed Below*** | | |
|  | **290.0** Senile Dementia, Uncomplicated, **290.2** Senile Dementia, with delusional or depressive features, **290.20** Senile Dementia, with Delusional Features, **290.21** Senile Dementia, with Depressive Features, **290.3** Senile Dementia, with Delirium |  |
|  | **294.2** Dementia, Unspecified, **294.20** Dementia, Unspecified Dementia, without Behavioral Disturbance, **294.21** Unspecified Dementia, with Behavioral Disturbance,**294.8** Other Persistent Mental Disorders | **F03.90** Unspecified dementia without behavioral disturbance, **F03.91** Unspecified Dementia, with behavioral disturbance |
| ***AD-RD = AD+ and Related Dementias (through Vascular Dementia below)*** | | |
|  | **331.1** Frontotemporal Dementia, **331.19** Other frontotemporal dementia, **331.2** Senile Degeneration of Brain, **331.82** Lewy Body Dementia | **G31.0** Frontotemporal Dementia, **G31.09** Other frontotemporal dementia, **G31.1** Senile Degeneration of Brain, Not Elsewhere Classified, G31.83 Lewy Body Dementia |
|  | **290.1** Presenile Dementia, **290.10** Presenile Dementia, uncomplicated, **290.11** Presenile Dementia, with delirium, **290.12** Presenile Dementia, with delusional features, **290.13** Presenile Dementia, with depressive features | **F03.90** Unspecified Dementia, without behavioral disturbance, **F05** Delirium due to known physiological condition |
|  | **331.5** Idiopathic Normal Pressure Hydrocephalus (INPH) | **G91.2** Idiopathic Normal Pressure Hydrocephalus (INPH) |
| ***Vascular Dementia (VaD)*** | | |
|  | **290.40** Vascular Dementia, Uncomplicated, **290.41** Vascular Dementia, with Delirium, **290.42** Vascular Dementia, with Delusions, **290.43** Vascular Dementia, with Depressed Mood | **F01.50** Vascular Dementia, without Behavioral Disturbance, **F01.51** Vascular Dementia, with Behavioral Disturbance |
| ***Non-specific Dementia not Mentioned Above*** | | |
|  | **294.1** Dementia in conditions classified elsewhere, **294.10** Dementia in conditions classified elsewhere without behavioral disturbance, **294.11** Dementia in conditions classified elsewhere with behavioral disturbance | **F02.80** Dementia in other diseases classified elsewhere without behavioral disturbance, **F02.81** Dementia in other diseases classified elsewhere with behavioral disturbance |
| ***Specific Dementia Not Mentioned Above*** | | |
|  | **333.4** Huntington’s Disease, **331.11** Pick’s Disease | **G10** Huntington’s Disease**, A81.00** Creutzfeldt-Jakob Disease, **G31.01** Pick's disease, **F10.96** Korsakoff Syndrome |

**Methods S1: Two-Sample MR Approaches**

To assess the robustness of the association and of the estimate results from the 2SLS approach, we also utilized MR methods created to analyze causal effects when conducting two-sample MR. The inverse-weighted (IVW) method uses the inverse of the standard errors of individual variants as weights in constructing the best fit line between the exposure effects and the outcome effects. IVW assumes there is no pleiotropy, forcing the line to go through the origin. The median method assumes that as many as half of the instruments could be invalid and uses the median as the effect estimate. MR Egger^1^ is an approach that not only tests for pleiotropy but also provides an estimate of the intercept and a p-value for the probability of its difference from zero while providing an estimate of the relationship between the instrument and the outcome. MR-PRESSO^2^ provides an effect estimate based on eliminating outliers from the score. For all these methods except MR-PRESSO, we utilized the MendelianRandomization^3^ R package v0.3.0 which provides a concise summary of estimates, confidence intervals and p-values as well as visual representations of the results. For MR-PRESSO, we used the R package version 1.0 from Verbanck et al.^2^ Note that when applying two-sample methods, we utilized the variants determined to be valid instruments from the Mahajan et. al., 2018 publication,^4^ for our effect sizes and standard errors for the exposure.

| **Statistic** | **p-value** | **Diagnosis** | **HARE** |
| --- | --- | --- | --- |
| -4.1319 | 3.60E-05 | All Cause Dementia | EUR |
| -0.2254 | 0.8216 | Clinical AD | EUR |
| -4.1325 | 3.59E-05 | Clinical VaD | EUR |
| -0.7742 | 0.4388 | All Cause Dementia | AFR |
| -0.4832 | 0.6289 | Clinical AD | AFR |
| -1.9078 | 0.0564 | Clinical VaD | AFR |
| 0.4536 | 0.6501 | All Cause Dementia | HIS |
| 1.6461 | 0.0997 | Clinical AD | HIS |
| -0.3257 | 0.7447 | Clinical VaD | HIS |

**Table S2.** Cochran-Armitage trend test results for the relationship between quartiles of the predicted probability of diabetes and dementia diagnoses

**Table S3.** MR estimates for all dementia diagnoses by HARE group. All_Cause refers to all-cause-dementia, VaD refers to clinically diagnosed VaD, AD refers to clinically diagnosed AD. Simple estimates have no weighting, weighted estimates use standard errors for its weighting factor, penalized estimates down-weight the role of outliers, and robust estimates utilize alternate techniques to typical linear regression, relaxing assumptions like heteroscedasticity and distribution of the error terms in the model.^1,3,5^

| **Method** | **OR** | **95% CI** | **p-value** | **Ancestry** | **Outcome** |
| --- | --- | --- | --- | --- | --- |
| Simple median | 1.11 | [1.067,1.159] | 4.68E-07 | EUR | All_Cause |
| Weighted median | 1.08 | [1.038,1.125] | 1.77E-04 | EUR | All_Cause |
| Penalized weighted median | 1.08 | [1.039,1.126] | 1.44E-04 | EUR | All_Cause |
| IVW | 1.09 | [1.062,1.117] | 3.39E-11 | EUR | All_Cause |
| Penalized IVW | 1.10 | [1.075,1.127] | 1.14E-15 | EUR | All_Cause |
| Robust IVW | 1.10 | [1.065,1.128] | 3.49E-10 | EUR | All_Cause |
| Penalized robust IVW | 1.10 | [1.070,1.132] | 3.74E-11 | EUR | All_Cause |
| MR-Egger | 1.08 | [1.032,1.133] | 9.69E-04 | EUR | All_Cause |
| (intercept) | 1.00 | [0.998,1.003] | 7.23E-01 | EUR | All_Cause |
| Penalized MR-Egger | 1.09 | [1.041,1.136] | 1.49E-04 | EUR | All_Cause |
| (intercept) | 1.00 | [0.998,1.003] | 5.83E-01 | EUR | All_Cause |
| Robust MR-Egger | 1.08 | [1.025,1.141] | 4.14E-03 | EUR | All_Cause |
| (intercept) | 1.00 | [0.998,1.004] | 5.43E-01 | EUR | All_Cause |
| Penalized robust MR-Egger | 1.08 | [1.026,1.142] | 3.52E-03 | EUR | All_Cause |
| (intercept) | 1.00 | [0.998,1.004] | 4.94E-01 | EUR | All_Cause |
| Simple median | 1.15 | [1.046,1.266] | 3.89E-03 | EUR | VaD |
| Weighted median | 1.10 | [0.985,1.218] | 9.34E-02 | EUR | VaD |
| Penalized weighted median | 1.10 | [0.985,1.218] | 9.26E-02 | EUR | VaD |
| IVW | 1.16 | [1.093,1.221] | 3.32E-07 | EUR | VaD |
| Penalized IVW | 1.15 | [1.092,1.220] | 3.60E-07 | EUR | VaD |
| Robust IVW | 1.16 | [1.094,1.226] | 4.35E-07 | EUR | VaD |
| Penalized robust IVW | 1.16 | [1.094,1.226] | 4.37E-07 | EUR | VaD |
| MR-Egger | 1.15 | [1.036,1.270] | 8.53E-03 | EUR | VaD |
| (intercept) | 1.00 | [0.994,1.007] | 8.71E-01 | EUR | VaD |
| Penalized MR-Egger | 1.16 | [1.044,1.280] | 5.40E-03 | EUR | VaD |
| (intercept) | 1.00 | [0.994,1.006] | 9.78E-01 | EUR | VaD |
| Robust MR-Egger | 1.15 | [1.029,1.284] | 1.34E-02 | EUR | VaD |
| (intercept) | 1.00 | [0.993,1.008] | 8.82E-01 | EUR | VaD |
| Penalized robust MR-Egger | 1.15 | [1.031,1.287] | 1.23E-02 | EUR | VaD |
| (intercept) | 1.00 | [0.993,1.008] | 9.12E-01 | EUR | VaD |
| Simple median | 1.08 | [0.990,1.185] | 8.00E-02 | EUR | AD |
| Weighted median | 1.01 | [0.917,1.114] | 8.31E-01 | EUR | AD |
| Penalized weighted median | 1.01 | [0.909,1.110] | 9.27E-01 | EUR | AD |
| IVW | 1.07 | [1.010,1.127] | 2.09E-02 | EUR | AD |
| Penalized IVW | 1.07 | [1.013,1.126] | 1.46E-02 | EUR | AD |
| Robust IVW | 1.07 | [1.011,1.125] | 1.78E-02 | EUR | AD |
| Penalized robust IVW | 1.07 | [1.012,1.125] | 1.65E-02 | EUR | AD |
| MR-Egger | 1.07 | [0.964,1.180] | 2.09E-01 | EUR | AD |
| (intercept) | 1.00 | [0.994,1.006] | 9.95E-01 | EUR | AD |
| Penalized MR-Egger | 1.06 | [0.966,1.173] | 2.10E-01 | EUR | AD |
| (intercept) | 1.00 | [0.994,1.006] | 9.40E-01 | EUR | AD |
| Robust MR-Egger | 1.05 | [0.960,1.150] | 2.84E-01 | EUR | AD |
| (intercept) | 1.00 | [0.995,1.008] | 7.27E-01 | EUR | AD |
| Penalized robust MR-Egger | 1.05 | [0.960,1.148] | 2.85E-01 | EUR | AD |
| (intercept) | 1.00 | [0.995,1.008] | 7.13E-01 | EUR | AD |
| Simple median | 1.12 | [1.003,1.256] | 4.45E-02 | AFR | All_Cause |
| Weighted median | 1.09 | [0.953,1.256] | 2.01E-01 | AFR | All_Cause |
| Penalized weighted median | 1.09 | [0.953,1.257] | 2.02E-01 | AFR | All_Cause |
| IVW | 1.07 | [1.002,1.141] | 4.25E-02 | AFR | All_Cause |
| Penalized IVW | 1.07 | [1.002,1.140] | 4.27E-02 | AFR | All_Cause |
| Robust IVW | 1.08 | [1.014,1.142] | 1.62E-02 | AFR | All_Cause |
| Penalized robust IVW | 1.08 | [1.014,1.142] | 1.62E-02 | AFR | All_Cause |
| MR-Egger | 0.98 | [0.872,1.110] | 7.89E-01 | AFR | All_Cause |
| (intercept) | 1.01 | [0.999,1.014] | 1.09E-01 | AFR | All_Cause |
| Penalized MR-Egger | 0.99 | [0.877,1.115] | 8.55E-01 | AFR | All_Cause |
| (intercept) | 1.01 | [0.998,1.013] | 1.34E-01 | AFR | All_Cause |
| Robust MR-Egger | 1.00 | [0.886,1.124] | 9.75E-01 | AFR | All_Cause |
| (intercept) | 1.01 | [0.998,1.014] | 1.50E-01 | AFR | All_Cause |
| Penalized robust MR-Egger | 1.00 | [0.888,1.125] | 9.96E-01 | AFR | All_Cause |
| (intercept) | 1.01 | [0.998,1.013] | 1.57E-01 | AFR | All_Cause |
| Simple median | 1.08 | [0.881,1.327] | 4.57E-01 | AFR | VaD |
| Weighted median | 1.25 | [0.975,1.607] | 7.80E-02 | AFR | VaD |
| Penalized weighted median | 1.25 | [0.975,1.607] | 7.82E-02 | AFR | VaD |
| IVW | 1.15 | [1.025,1.297] | 1.74E-02 | AFR | VaD |
| Penalized IVW | 1.16 | [1.028,1.301] | 1.54E-02 | AFR | VaD |
| Robust IVW | 1.15 | [1.023,1.297] | 1.97E-02 | AFR | VaD |
| Penalized robust IVW | 1.15 | [1.024,1.296] | 1.88E-02 | AFR | VaD |
| MR-Egger | 1.20 | [0.967,1.499] | 9.64E-02 | AFR | VaD |
| (intercept) | 1.00 | [0.983,1.010] | 6.45E-01 | AFR | VaD |
| Penalized MR-Egger | 1.21 | [0.969,1.502] | 9.32E-02 | AFR | VaD |
| (intercept) | 1.00 | [0.984,1.011] | 6.81E-01 | AFR | VaD |
| Robust MR-Egger | 1.22 | [1.022,1.446] | 2.76E-02 | AFR | VaD |
| (intercept) | 1.00 | [0.983,1.009] | 5.22E-01 | AFR | VaD |
| Penalized robust MR-Egger | 1.22 | [1.022,1.446] | 2.73E-02 | AFR | VaD |
| (intercept) | 1.00 | [0.983,1.009] | 5.26E-01 | AFR | VaD |
| Simple median | 1.37 | [1.045,1.794] | 2.27E-02 | AFR | AD |
| Weighted median | 1.45 | [1.103,1.895] | 7.60E-03 | AFR | AD |
| Penalized weighted median | 1.47 | [1.097,1.976] | 9.97E-03 | AFR | AD |
| IVW | 1.14 | [0.973,1.329] | 1.06E-01 | AFR | AD |
| Penalized IVW | 1.14 | [0.978,1.331] | 9.44E-02 | AFR | AD |
| Robust IVW | 1.16 | [0.974,1.386] | 9.63E-02 | AFR | AD |
| Penalized robust IVW | 1.16 | [0.975,1.386] | 9.29E-02 | AFR | AD |
| MR-Egger | 1.05 | [0.781,1.398] | 7.68E-01 | AFR | AD |
| (intercept) | 1.01 | [0.988,1.025] | 4.99E-01 | AFR | AD |
| Penalized MR-Egger | 1.05 | [0.789,1.406] | 7.27E-01 | AFR | AD |
| (intercept) | 1.01 | [0.988,1.024] | 5.31E-01 | AFR | AD |
| Robust MR-Egger | 1.08 | [0.699,1.659] | 7.37E-01 | AFR | AD |
| (intercept) | 1.01 | [0.982,1.030] | 6.61E-01 | AFR | AD |
| Penalized robust MR-Egger | 1.08 | [0.703,1.658] | 7.27E-01 | AFR | AD |
| (intercept) | 1.01 | [0.982,1.029] | 6.69E-01 | AFR | AD |
| Simple median | 1.08 | [0.927,1.261] | 3.21E-01 | HIS | All_Cause |
| Weighted median | 1.16 | [0.965,1.384] | 1.17E-01 | HIS | All_Cause |
| Penalized weighted median | 1.16 | [0.964,1.385] | 1.17E-01 | HIS | All_Cause |
| IVW | 1.08 | [0.986,1.188] | 9.49E-02 | HIS | All_Cause |
| Penalized IVW | 1.08 | [0.986,1.183] | 9.58E-02 | HIS | All_Cause |
| Robust IVW | 1.08 | [0.998,1.178] | 5.44E-02 | HIS | All_Cause |
| Penalized robust IVW | 1.08 | [0.998,1.176] | 5.50E-02 | HIS | All_Cause |
| MR-Egger | 1.12 | [0.937,1.329] | 2.18E-01 | HIS | All_Cause |
| (intercept) | 1.00 | [0.987,1.009] | 6.84E-01 | HIS | All_Cause |
| Penalized MR-Egger | 1.12 | [0.946,1.329] | 1.87E-01 | HIS | All_Cause |
| (intercept) | 1.00 | [0.987,1.008] | 6.07E-01 | HIS | All_Cause |
| Robust MR-Egger | 1.12 | [0.983,1.275] | 9.00E-02 | HIS | All_Cause |
| (intercept) | 1.00 | [0.988,1.008] | 6.33E-01 | HIS | All_Cause |
| Penalized robust MR-Egger | 1.12 | [0.986,1.276] | 8.19E-02 | HIS | All_Cause |
| (intercept) | 1.00 | [0.988,1.007] | 5.99E-01 | HIS | All_Cause |
| Simple median | 1.32 | [0.957,1.831] | 9.03E-02 | HIS | VaD |
| Weighted median | 0.97 | [0.705,1.320] | 8.22E-01 | HIS | VaD |
| Penalized weighted median | 0.94 | [0.686,1.292] | 7.09E-01 | HIS | VaD |
| IVW | 1.06 | [0.876,1.283] | 5.49E-01 | HIS | VaD |
| Penalized IVW | 1.08 | [0.892,1.306] | 4.31E-01 | HIS | VaD |
| Robust IVW | 1.08 | [0.871,1.343] | 4.79E-01 | HIS | VaD |
| Penalized robust IVW | 1.09 | [0.877,1.349] | 4.43E-01 | HIS | VaD |
| MR-Egger | 0.91 | [0.633,1.304] | 6.04E-01 | HIS | VaD |
| (intercept) | 1.01 | [0.989,1.034] | 3.25E-01 | HIS | VaD |
| Penalized MR-Egger | 0.89 | [0.623,1.283] | 5.45E-01 | HIS | VaD |
| (intercept) | 1.01 | [0.991,1.037] | 2.34E-01 | HIS | VaD |
| Robust MR-Egger | 0.87 | [0.581,1.294] | 4.85E-01 | HIS | VaD |
| (intercept) | 1.02 | [0.992,1.041] | 2.03E-01 | HIS | VaD |
| Penalized robust MR-Egger | 0.86 | [0.582,1.283] | 4.68E-01 | HIS | VaD |
| (intercept) | 1.02 | [0.993,1.041] | 1.77E-01 | HIS | VaD |
| Simple median | 0.99 | [0.701,1.385] | 9.33E-01 | HIS | AD |
| Weighted median | 0.93 | [0.623,1.386] | 7.18E-01 | HIS | AD |
| Penalized weighted median | 0.93 | [0.619,1.387] | 7.13E-01 | HIS | AD |
| IVW | 1.02 | [0.832,1.238] | 8.82E-01 | HIS | AD |
| Penalized IVW | 1.00 | [0.821,1.217] | 9.99E-01 | HIS | AD |
| Robust IVW | 1.00 | [0.820,1.207] | 9.57E-01 | HIS | AD |
| Penalized robust IVW | 0.99 | [0.819,1.201] | 9.33E-01 | HIS | AD |
| MR-Egger | 0.97 | [0.666,1.406] | 8.64E-01 | HIS | AD |
| (intercept) | 1.00 | [0.980,1.027] | 7.67E-01 | HIS | AD |
| Penalized MR-Egger | 0.95 | [0.654,1.371] | 7.71E-01 | HIS | AD |
| (intercept) | 1.00 | [0.981,1.027] | 7.30E-01 | HIS | AD |
| Robust MR-Egger | 0.92 | [0.679,1.241] | 5.79E-01 | HIS | AD |
| (intercept) | 1.01 | [0.986,1.027] | 5.61E-01 | HIS | AD |
| Penalized robust MR-Egger | 0.92 | [0.679,1.233] | 5.60E-01 | HIS | AD |
| (intercept) | 1.01 | [0.986,1.027] | 5.56E-01 | HIS | AD |

**Results S1: Two-Sample MR Results**

The results for the association between GRS330 and VaD using two sample methods were directionally consistent with the primary one-sample analysis for EUR (Figure S2, top left) and AFR (Figure S2, middle left) while the EUR results, generally met significance thresholds and the AFR results did not (Table S3). The results for HIS did not show a consistent association of GRS330 with VaD using two sample MR methods (Figure S2, bottom left, eTable4). The results for the associations between GRS330 and AD only reached statistical significance in the weighted median method for AFR, (Table S3) but otherwise showed a consistent direction of effect for EUR and AFR (Figure S2, top right and middle right, respectively). The HIS results were not significant (Table S3) and appeared to be protective (Figure S2, bottom right). MR_PRESSO results (Table S4), when using the ‘outlier-corrected’ estimate consistently showed an increased effect in the same range as the MendelianRandomization package estimates.

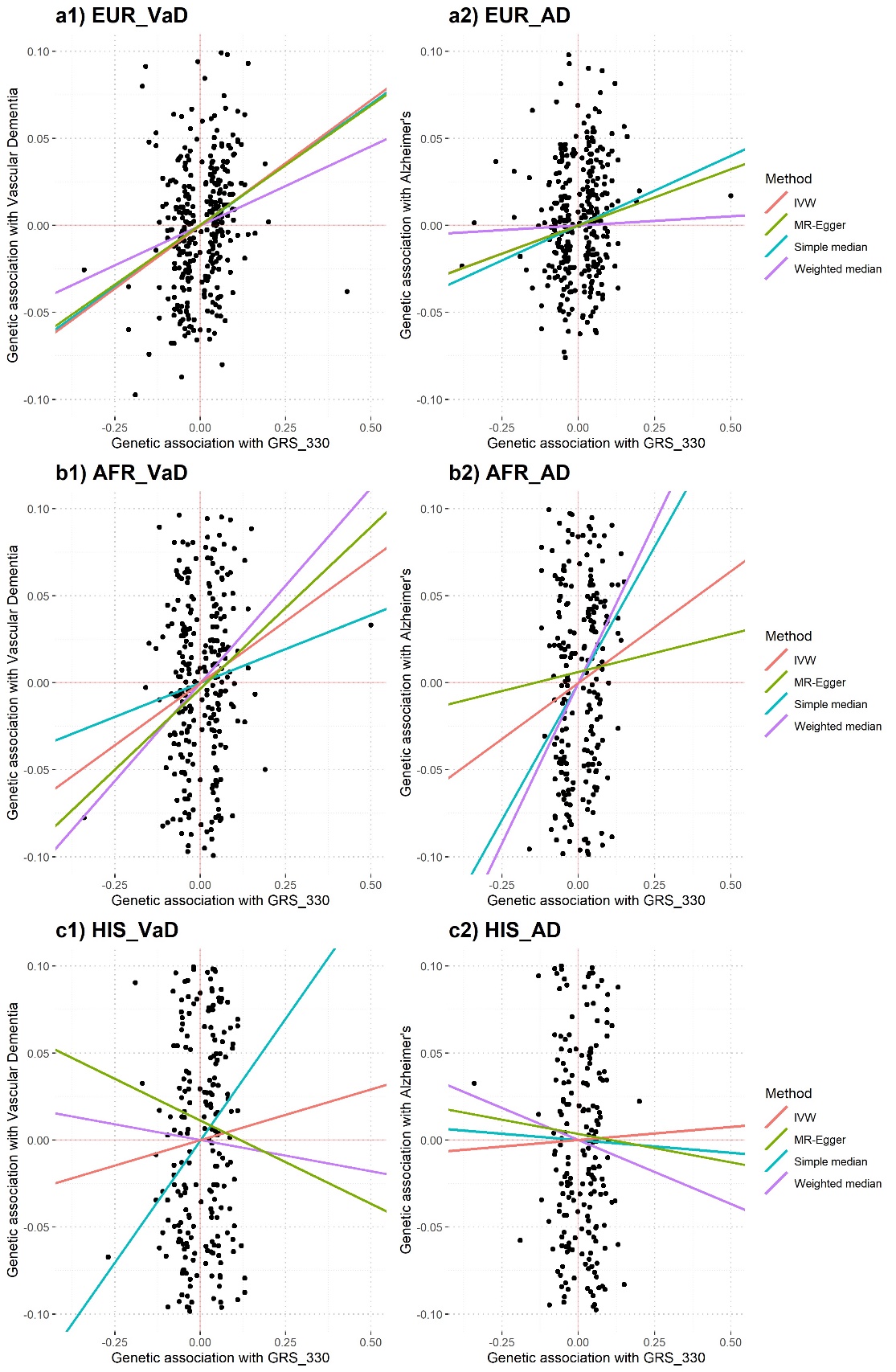

**Figure S2.** Methods using the Mendelian Randomization package are shown here. Diagrams are shown by HARE group: Top Row: European, Middle Row: AFR: African, Bottom Row: HIS: Hispanic and clinical diagnosis: VaD for vascular dementia, AD for Alzheimer’s disease. Simple estimates have no weighting and weighted estimates use standard errors for its weighting factor.^1,3,5^ Note that the IVW line is completely covered by the MR Egger line in EUR_AD (Top, Right) and so, is not visible.

**Table S4.** Pleiotropy assumption – Using MR Egger^3^

| **Outcome** | **Estimate** | **Causal.pval** | **Intercept** | **Pleio.pval** | **SNPs** | **Heterogeneity p-value** | **Ancestry** |
| --- | --- | --- | --- | --- | --- | --- | --- |
| All_Cause | 0.0673 | 0.2123 | -0.0005 | 0.8930 | 331 | 4.33E-240 | EUR |
| All_Cause | 0.0784 | 0.0010 | 0.0005 | 0.7231 | 330 | 0.0103 | EUR |
| StrictAD | 0.0401 | 0.7165 | -0.0022 | 0.7523 | 331 | 4.92E-178 | EUR |
| StrictAD | 0.0648 | 0.2089 | 0.0000 | 0.9947 | 330 | 0.0880 | EUR |
| VaD | 0.1224 | 0.1271 | -0.0007 | 0.8818 | 331 | 2.35E-39 | EUR |
| VaD | 0.1371 | 0.0085 | 0.0005 | 0.8706 | 330 | 0.4511 | EUR |
| All_Cause | -0.0297 | 0.6965 | 0.0051 | 0.2788 | 331 | 8.32E-10 | AFR |
| All_Cause | -0.0165 | 0.7888 | 0.0061 | 0.1089 | 330 | 0.4470 | AFR |
| StrictAD | 0.0103 | 0.9554 | 0.0036 | 0.7503 | 331 | 1.47E-12 | AFR |
| StrictAD | 0.0437 | 0.7684 | 0.0063 | 0.4985 | 330 | 0.1626 | AFR |
| VaD | 0.1692 | 0.1660 | -0.0045 | 0.5565 | 331 | 8.91E-03 | AFR |
| VaD | 0.1859 | 0.0964 | -0.0032 | 0.6453 | 330 | 0.7262 | AFR |
| All_Cause | 0.1022 | 0.2840 | -0.0028 | 0.6337 | 331 | 3.58E-03 | HIS |
| All_Cause | 0.1097 | 0.2176 | -0.0022 | 0.6845 | 330 | 0.2045 | HIS |
| StrictAD | -0.0514 | 0.8034 | 0.0020 | 0.8758 | 331 | 3.47E-03 | HIS |
| StrictAD | -0.0327 | 0.8637 | 0.0035 | 0.7671 | 330 | 0.2884 | HIS |
| VaD | -0.1067 | 0.5746 | 0.0104 | 0.3749 | 331 | 0.1787 | HIS |
| VaD | -0.0958 | 0.6035 | 0.0112 | 0.3252 | 330 | 0.4465 | HIS |

**Table S5.** MR_Presso^2^ Results when conducting an Outlier test using 10,000 iterations and a significance threshold of 0.05. Note that no outliers were found in HIS with this number of iterations.

| **MR Analysis** | **OR** | **95% CI** | **p-value** | **Population** | **Diagnosis** |
| --- | --- | --- | --- | --- | --- |
| Raw | 1.06 | [1,1.13] | 3.71E-02 | EUR | All_Cause |
| Outlier-corrected | 1.09 | [1.06,1.12] | 1.39E-10 | EUR | All_Cause |
| Raw | 1.01 | [0.9,1.14] | 8.57E-01 | EUR | StrictAD |
| Outlier-corrected | 1.07 | [1.01,1.13] | 2.15E-02 | EUR | StrictAD |
| Raw | 1.12 | [1.03,1.22] | 1.01E-02 | EUR | VaD |
| Outlier-corrected | 1.16 | [1.09,1.22] | 5.63E-07 | EUR | VaD |
| Raw | 1.04 | [0.96,1.13] | 3.25E-01 | AFR | All_Cause |
| Outlier-corrected | 1.07 | [1,1.14] | 4.33E-02 | AFR | All_Cause |
| Raw | 1.06 | [0.88,1.29] | 5.44E-01 | AFR | StrictAD |
| Outlier-corrected | 1.14 | [0.97,1.33] | 1.07E-01 | AFR | StrictAD |
| Raw | 1.11 | [0.98,1.27] | 9.73E-02 | AFR | VaD |
| Outlier-corrected | 1.15 | [1.03,1.29] | 1.52E-02 | AFR | VaD |
| Raw | 1.07 | [0.96,1.18] | 2.10E-01 | HIS | All_Cause |
| Outlier-corrected | 1.08 | [0.99,1.19] | 9.59E-02 | HIS | All_Cause |
| Raw | 0.98 | [0.79,1.21] | 8.27E-01 | HIS | StrictAD |
| Outlier-corrected | 1.02 | [0.83,1.24] | 8.82E-01 | HIS | StrictAD |
| Raw | 1.04 | [0.85,1.26] | 7.15E-01 | HIS | VaD |
| Outlier-corrected | NA | [NA,NA] | NA | HIS | VaD |

**Table S6.** Sensitivity E-value^6^: The strength of unmeasured confounding required to completely attenuate the relationship between the exposure (diabetes) and the outcome (dementia). The table is sorted in order of decreasing E-Values.

| **Outcome** | **Ancestry** | **OR [95% CI]** | **E-Value of Estimate** | **E-Value of Lower CI** | **p.value** |
| --- | --- | --- | --- | --- | --- |
| Vascular_Dementia | AFR | 1.15 [1.03,1.3] | 1.57 | 1.21 | 0.0159 |
| Vascular_Dementia | EUR | 1.14 [1.09,1.21] | 1.54 | 1.40 | 7.20E-07 |
| Strict_AD | AFR | 1.14 [0.98,1.32] | 1.54 | 1.00 | 0.0958 |
| All_Cause_Dementia | EUR | 1.08 [1.06,1.11] | 1.37 | 1.31 | 2.64E-12 |
| All_Cause_Dementia | HIS | 1.08 [0.99,1.18] | 1.37 | 1.00 | 0.0926 |
| All_Cause_Dementia | AFR | 1.07 [1,1.14] | 1.34 | 1.00 | 0.0419 |
| Strict_AD | EUR | 1.06 [1.01,1.12] | 1.31 | 1.11 | 0.0176 |
| Vascular_Dementia | HIS | 1.06 [0.88,1.28] | 1.31 | 1.00 | 0.5307 |
| Strict_AD | HIS | 1.02 [0.84,1.23] | 1.16 | 1.00 | 0.8743 |

**Table S7.** Table of results when adjusting for BMI in Stage 2 of the two stage least squares method

| **Covariate** | **OR [95% CI]** | **p-value** | **Diagnosis** | **HARE** |
| --- | --- | --- | --- | --- |
| (Quart)(-0.873,-0.342] | 1.06 [1.02,1.11] | 5.30E-03 | All Cause Dementia | EUR |
| (Quart)(-0.342,0.282] | 1.1 [1.05,1.15] | 1.40E-05 | All Cause Dementia | EUR |
| (Quart)(0.282,4.76] | 1.18 [1.13,1.23] | 6.30E-14 | All Cause Dementia | EUR |
| bmi | 0.99 [0.98,0.99] | 1.20E-19 | All Cause Dementia | EUR |
| (Quart)(-0.873,-0.342] | 1.16 [1.04,1.28] | 7.40E-03 | Clinical VaD | EUR |
| (Quart)(-0.342,0.282] | 1.22 [1.1,1.36] | 1.90E-04 | Clinical VaD | EUR |
| (Quart)(0.282,4.76] | 1.33 [1.2,1.48] | 8.30E-08 | Clinical VaD | EUR |
| bmi | 1 [0.99,1] | 5.00E-01 | Clinical VaD | EUR |
| (Quart)(-0.873,-0.342] | 1.09 [0.99,1.2] | 9.00E-02 | Clinical AD | EUR |
| (Quart)(-0.342,0.282] | 1.08 [0.98,1.19] | 1.20E-01 | Clinical AD | EUR |
| (Quart)(0.282,4.76] | 1.13 [1.02,1.25] | 1.40E-02 | Clinical AD | EUR |
| bmi | 0.96 [0.96,0.97] | 4.60E-21 | Clinical AD | EUR |
| (Quart)(0.578,1.02] | 1.15 [1.02,1.28] | 1.70E-02 | All Cause Dementia | AFR |
| (Quart)(1.02,1.5] | 1.1 [0.98,1.23] | 1.00E-01 | All Cause Dementia | AFR |
| (Quart)(1.5,4.28] | 1.17 [1.05,1.31] | 6.20E-03 | All Cause Dementia | AFR |
| bmi | 0.97 [0.97,0.98] | 1.20E-11 | All Cause Dementia | AFR |
| (Quart)(0.578,1.02] | 1.38 [1.13,1.7] | 2.00E-03 | Clinical VaD | AFR |
| (Quart)(1.02,1.5] | 1.17 [0.95,1.45] | 1.40E-01 | Clinical VaD | AFR |
| (Quart)(1.5,4.28] | 1.45 [1.18,1.78] | 4.30E-04 | Clinical VaD | AFR |
| bmi | 1 [0.99,1.01] | 8.00E-01 | Clinical VaD | AFR |
| (Quart)(0.578,1.02] | 1.1 [0.84,1.44] | 4.90E-01 | Clinical AD | AFR |
| (Quart)(1.02,1.5] | 1.13 [0.87,1.48] | 3.70E-01 | Clinical AD | AFR |
| (Quart)(1.5,4.28] | 1.28 [0.99,1.67] | 6.20E-02 | Clinical AD | AFR |
| bmi | 0.93 [0.91,0.95] | 3.90E-12 | Clinical AD | AFR |
| (Quart)(0.367,1.08] | 0.98 [0.83,1.16] | 8.20E-01 | All Cause Dementia | HIS |
| (Quart)(1.08,1.87] | 1.14 [0.97,1.34] | 1.20E-01 | All Cause Dementia | HIS |
| (Quart)(1.87,5.61] | 1.02 [0.86,1.21] | 8.20E-01 | All Cause Dementia | HIS |
| bmi | 0.97 [0.96,0.99] | 6.50E-05 | All Cause Dementia | HIS |
| (Quart)(0.367,1.08] | 1.12 [0.78,1.59] | 5.40E-01 | Clinical VaD | HIS |
| (Quart)(1.08,1.87] | 1.06 [0.74,1.53] | 7.40E-01 | Clinical VaD | HIS |
| (Quart)(1.87,5.61] | 1.27 [0.9,1.81] | 1.70E-01 | Clinical VaD | HIS |
| bmi | 0.99 [0.97,1.02] | 6.80E-01 | Clinical VaD | HIS |
| (Quart)(0.367,1.08] | 0.88 [0.62,1.25] | 4.70E-01 | Clinical AD | HIS |
| (Quart)(1.08,1.87] | 0.98 [0.7,1.39] | 9.30E-01 | Clinical AD | HIS |
| (Quart)(1.87,5.61] | 0.77 [0.53,1.12] | 1.70E-01 | Clinical AD | HIS |
| bmi | 0.94 [0.91,0.97] | 8.50E-05 | Clinical AD | HIS |
